# Impact of ‘never use’ abbreviations (Error Prone Abbreviations-EPA’s) list on incidence of EPAs in inpatient medical prescriptions in apex tertiary care public hospital in India

**DOI:** 10.1101/2020.10.05.20206896

**Authors:** Arif Thachaparambil, Mohammad Kausar, R Mahesh, DK Sharma

## Abstract

**Background:** Abbreviations are commonly used in medical records to save time and space but use in prescriptions can be a reason for communication failures and preventable harm during healthcare delivery. Nearly 5% of medication errors can be attributable to abbreviation use. Prescriptions need to be clear so that nurses and pharmacists can correctly interpret intentions of doctors. For patient safety, hospitals should implement a process for uniform use of approved abbreviations, such as through use of an approved list or never-use list of abbreviations and symbols.

**Local Problem:** In the hospital under study, there was no system of avoiding error prone abbreviations while prescribing to prevent medical errors. Hence, an interventional study was done to quantify and reduce incidence of error prone abbreviations.

**Methods:** The study design was pre-post interventional / quasi-experimental design to assess the impact of never use list and standardized abbreviations. The study was conducted after ethical approval from Institute Ethics Committee. Pre intervention data was collected by a retrospective closed in-patient medical record review. Post interventional incidence of error prone abbreviations was determined and the effectiveness of the same was assessed by using statistical analysis.

**Intervention:** An approved ‘Never use’ list and standardized abbreviations were developed and poster copies were affixed in inpatient wards, doctors were educated and poster pamphlets were also distributed.

**Results:** Incidence of error abbreviations in inpatient prescription were 47.5% and ‘Never Use’ list of abbreviation led to a statistically significant reduction of error-prone abbreviation by 8.2% from 47.5% to 43.6% (P\0.006)

**Conclusion:** ‘Never Use’ lists are effective in reducing incidence of common error-prone abbreviations and discipline wise variation is observed. Adoption of such lists is highly recommended. The lists should be comprehensive, regularly updated and educational interventions should be comprehensive and integration into patient medical charts and pocket friendly flash cards may be provided for better outcomes. Enforcing a policy to prohibit the use of EPAs while prescribing will also be helpful.

## INTRODUCTION

### Problem Description

Abbreviations are commonly used in medical records to save time and space. However, abbreviations can be confusing and may convey different meaning in different contexts, e.g., DOA is interpreted as either ‘date of admission’ or ‘dead on arrival’ by various clinical or surgical disciplines. Good medical practice warrants that doctors keep clear, accurate and legible medical records to provide good clinical care and enable correct interpretation by pharmacists and nurses. Inappropriate abbreviations in prescriptions may alter intended therapeutic outcomes and even harm patients. [1] Communication failures often result from non-uniform or non-standardized use of abbreviations, symbols, and codes and cause patient safety incidents. [2] Nearly 5% of errors reported were attributable to abbreviation use. [3]

### Available knowledge

The problem of inappropriate use of abbreviations has been raised by many medication safety organizations such as National Coordinating Council for Medication Error Reporting and Prevention of the USA, Institute of Safe Medication Practices, etc. National Board for Accreditation of Hospitals and Healthcare providers (NABH) quality standards recommends standardized list of approved and documented abbreviations for medication orders for uniform use throughout to ensure that the abbreviation has only one meaning thereby avoiding medical errors. Dangerous/ error prone abbreviations (EPA’s) should not be used. The standardized approved list of abbreviations, symbols, and dose designations should be based on best national and international practices. [4] Joint Commission International (JCI) accreditation standards recommend developing and/or adopt a do-not-use list of abbreviations and symbols. [2] EPAs and correct way of abbreviations are also recommended by ISMP (Institute for Safe Medication Practices) and ACSQH (Australian Commission on Safety and Quality in Health Care). [5]

### Rationale

In the hospital under study, there was no system of avoiding error prone abbreviations while prescribing to prevent medical errors.

### Specific Aims

An interventional study was done to quantify and reduce incidence of error prone abbreviations.

## METHODS

### Context

In the hospital under study, being a multi-specialty hospital, there were many specialty inpatient wards. Being hospital administrators, the researchers were aware that there was no formal mechanism in place to address potential harm by error prone abbreviations. Since broad specialty admitted most of the patients, the intervention was envisaged in these wards. The team involved in the work comprised of hospital administrators who were administrative in charges of the inpatient wards expected to ensure smooth and safe hospital operations related to patient care.

### Study of the Intervention(s)

Review of literature had indicated that many medication safety organisations had suggested use of standardized list of approved and documented abbreviations for medication orders. Use of do-not-use lists has also been recommended. Hence, an intervention of introducing never use list was decided.

To ensure the effect establish whether the observed outcomes were due to the intervention, pre-post study of prescriptions of the same inpatient areas was conducted and results compared.

### Study Design

The study design was pre-post interventional study / quasi-experimental study design to assess the impact of never use list and standardised abbreviations.

### Study Settings

This prospective study was conducted in inpatient wards of broad specialties of medicine, paediatrics and surgery of a tertiary care hospital.

### Study Period

The duration of this study was four months from July to November 2019. The objectives were to develop and implement a standardised “never use” list of error prone abbreviations and standardised abbreviations based on international bets practices and impact its list on incidence of prescription of error-prone abbreviations. The study was conducted after ethical approval from Institute Ethics Committee (IECPG-196).

### Sample Size

A pilot study showed that incidence of error-prone abbreviations in prescriptions varied from 8% to 40 % in broad specialties studied. One thousand three hundred prescriptions were studied - 600 during pre-interventional phase and 700 prescriptions in post interventional phase to identify a 10% change (before and after studies) in use of error-prone abbreviations in prescriptions taking 5% margin of error, 95% confidence in correlation with similar study in Hong Kong study which was done by Nithushi R, Samaranayake & Dixon S.

### Measures, Methodology and Intervention

Initially, a retrospective closed in-patient medical record review of patients from disciplines of medicine, paediatric and surgery was conducted using stratified random sampling to determine the incidence of these error prone abbreviations compared against lists by ISMP and ACSQH. Based on results of the pilot study, existing literature and inputs from experts in hospital administrations and patient safety, an approved ‘Never use’ list and standardised abbreviations were developed in the form of poster. Copies of these posters were affixed in inpatient wards of medicine, surgery and paediatric discipline, doctors were educated about EPAs and correct method of prescribing and poster pamphlets were also distributed. The post intervention data collection for calculating incident of error prone abbreviations in prescriptions was started two weeks after intervention.

### Statistical Analysis

Impact was assessed by comparing pre and post interventional results by data analysis using SPSS version 20. Paired t-test was applied to evaluate the statistical significance between pre and post interventional results.

### Ethical Considerations

The study was conducted after ethical approval from Institute Ethics Committee (IECPG-196).

## RESULTS

### Never use list of error prone abbreviations (EPAs)

By comparing the different international safety organisation, a list of 12 most error prone abbreviations was adopted which was frequently used in our hospital set up and the same list was considered as **“Never use”** list of error prone abbreviations (Table 1).

**Table 1.** “Never use” list of error prone abbreviations (EPAs) and correct prescribing.

| Sl. No | EPAs -ISMP /ACSQH | Intended meaning | Misinterpretation |
| --- | --- | --- | --- |
| 1. | INJ/ IJ | Injection | Mistaken as “IV” |
| 2. | Mg | Milligram | microgram |
| 3. | ug | Microgram | Mistaken as “mg” |
| 4. | gm | Gram | mg |
| 5. | od or OD | Once daily | right eye (OD-oculus dexter) |
| 6. | BID or BD/bd | Twice daily | bt-bed time |
| 7. | QID/qid | Four times daily | Mistaken as q1d(daily) |
| 8. | BT/bt | Bed time | bd-twice a day |
| 9. | Hs | At bed time | Half strength |
| 10. | IU | International units | IV (intravenous) |
| 11. | S/C | Subcutaneous | SL (sublingual) |
| 12. | Po/per os | Per oral | Left eye (OS- Oculus sinister) |

### Impact on incidence of EPAs

Among the 678 prescriptions that were selected for review, 78 prescriptions were excluded from the analysis because they contained non-drug items only. We reviewed 600 prescriptions in Phase II (post interventional phase), comprising a total of 2950 drug items.

### Pre-Intervention Incidence of EPAs

Pre intervention incidence of error prone abbreviation was higher in prescription of discipline of surgery (66%) followed by paediatrics (54.8%) and medicine (53.4%). Top three EPAs were ‘Inj’ instead of Injection (17%), followed by ‘BD’ instead of twice a day (10.8%) and OD instead of once daily (9.6%). (Table 2)

**Table 2.** Mean pre post interventional EPA incidence.

| EPA | Medicine |  | Surgery |  | Paediatrics |  | Total Incidence |  |  |  |
| --- | --- | --- | --- | --- | --- | --- | --- | --- | --- | --- |
|  | Pre<br>n=320 | Post<br>n=268 | Pre<br>n=385 | Post<br>n=355 | Pre<br>n=327 | Post<br>n=324 | Pre<br>n=1032 | Post<br>n=947 | Mean<br>Diff. | P-Value |
| **INJ/ IJ** | 10.5 | 9.8 | 23.6 | 22.8 | 16.8 | 16.3 | 17.0 | 16.3 | 0.7 | 0.011* |
| **Mg** | 2.1 | 1.7 | 0.3 | 1.0 | 0.6 | 0.4 | 1.0 | 1.0 | 0 | 0.893 |
| **ug** | 1.7 | 1.0 | 0.9 | 0.7 | 2.1 | 1.6 | 1.6 | 1.1 | 0.5 | 0.019* |
| **gm** | 2.0 | 1.0 | 3.5 | 3.6 | 1.7 | 2.0 | 2.4 | 2.2 | 0.2 | 0.635 |
| **od or OD** | 10.0 | 10.7 | 6.1 | 5.6 | 12.7 | 12.0 | 9.6 | 9.4 | 0.2 | 0.611 |
| **BID or BD/bd** | 11.0 | 9.2 | 12.7 | 11.9 | 8.8 | 8.0 | 10.8 | 9.7 | 1.1 | 0.093 |
| **QID/qid** | 1.6 | 0.8 | 4.3 | 1.0 | 0.9 | 2.1 | 2.3 | 1.3 | 1.0 | 0.556 |
| **Hs** | 1.0 | 0.9 | 1.2 | 1.9 | 0.3 | 1.0 | 0.8 | 1.3 | -0.5 | 0.296 |
| **IU** | 2.6 | 2.0 | 1.8 | 1.3 | 1.7 | 0.6 | 2.0 | 1.3 | 0.7 | 0.012* |
| **Dept wise. Total** | 42.5 | 37.1* | 54.4 | 49.8* | 45.6 | 44 | 47.5 | 43.6 | 3.9 | 0.006* |

### Post-Intervention Incidence of EPAs

Analysis of pre and post interventional results revealed statistically significant reduction 8.2% in overall use of EPAs from 47.5% to 43.6% (P\0.006) (Paired t-test).

### Discipline-wise impact on incidence of EPAs

Disciplines of medicine (15.9% reduction, p=0.024) and surgery (9.5% reduction, P=0.010) witnessed statistically significant reductions while there was no change in the discipline of paediatrics (1.5% reduction, P=0.81). (Table 2)

## DISCUSSION

### Key findings and Interpretations

The use of inappropriate abbreviations in prescriptions may alter intended therapeutic outcomes and even cause unnecessary harm. [5] Use of inappropriate abbreviations is one of the reasons for medication error. Hospitals opting for accreditation have preventive measures such as use of error prone abbreviations or do not use lists to guide prescribing behaviours. Others lack such initiatives. In Sudan, only 5% hospitals among 41 Khartoum State hospitals had a list of error-prone abbreviations. [6] A survey in the Pakistan established knowledge gap among medicine trainees regarding meaning and usage of common medical abbreviations where 93% used ‘IU’ instead of entire phrase ‘international unit’. [7]

Use of error prone abbreviations is pervasive but it incidence rates vary. In a multi-hospital Australian study hospitals, 76.9% patients had one or more errorLJprone abbreviations used in prescribing, with 8.4% of orders containing at least one errorLJprone abbreviation and 29.6% of these considered to be high risk for causing significant harm. [8] In another study in Australia, paediatric prescription audits identified error-prone abbreviations on 5% of audited medication orders. These were Qd, OD, U, mcg, trailing zeros or failure to include a leading zero when the dose is less than a one. [9] Audits in a Sri Lankan hospital, showed use of following error-prone abbreviations - μg (microgram), mcg (microgram), u (units), cc (cubic centimeter), OD (once a day), @ sign, d (days/daily), m (morning) and n (night) - at a rate of 17.4%, 0.1%, 1.9%, 0.2%, 0.2%, 4.9%, 23.5%, 4.4% and 15.8% respectively among all prescriptions reviewed. [10] In our study, overall incidence of error prone abbreviations before intervention was 47.5%. Top three EPAs were ‘Inj’ instead of Injection (17%), followed by ‘BD’ instead of twice a day (10.8%) and OD instead of once daily (9.6%). Common abbreviations recorded in acute care settings in Saudi Arabia were ‘IJ for injection’ (28.6%), ‘SC for subcutaneous’ (17.4%) and ‘OD for once daily’ (5.8%). [11] In another study in Riyadh, Saudi Arabia, use of unsafe abbreviations in 7000 medication orders was 28.3%. These abbreviations were used mainly by interns (41%), residents (33.6%), registrars (22%), and a few consultants (3.2%). The most common abbreviations used were ‘‘cc’’ instead of ‘‘mL’’ (50%), ‘‘@’’ instead of ‘‘at’’ (34%), and ‘‘DC’’ instead of ‘‘discontinue’’ (32%). [12] It appears that globally, the incidence somewhat inversely correlates with the socio-economic development of the countries.

In our study, introduction of never-use lists and standardised abbreviations have been effective in reducing incidence of EPA’s in prescriptions by 8.2% from 58.1% to 52.9%. In a similar pharmacist-led educational interventional study in Saudi Arabia, overall incidence of high risk abbreviations was reduced by 52% (53.6% to 25.5%). [11] The educational intervention was more comprehensive compared to that in our study. It included insertion of a printed list of abbreviations on brightly coloured paper into medical records/patient charts, pocket-sized flash cards provided for staff, laminated copies of list was attached to the back of the physician’s order divider in medical records. [11] These were in addition to the interventions undertaken in our study. A two phased intervention based on education and a hospital-wide policy on prohibition on use of unsafe abbreviations yielded 65% reduction from a baseline of 28.3%. [12] In our study, reductions in incidents of individual EPA’s was INJ/ IJ (4.1%), ug (31.3%) and IU (35%) study compared to reductions in EPAs in another similar study in Hong Kong on effectiveness of a ‘Do Not Use’ list that were 35.7% for ‘ug’, 92% for ‘od or OD’ and 69.2% for ‘IU’. [1]

It has been suggested introduction of such lists was effective but success largely depends a comprehensive EPA list. [13] Historically, these abbreviations crop up with alarming frequency. [14] Furthermore, frequent review and update of ‘‘Never Use’ list and frequent reminders recommended are recommended but absolute elimination may be difficult. Interventional study on use of electronic health records (EHR) technology has helped eliminate error prone abbreviation from a pre-intervention incidence rate of 41% to 0%. [15] However, implement of EHRs is difficult as it not only involves costly technology, its maintenance and training but in the fundamental way how a doctor behaves while prescribing. It is quite different from scribbling notes possible even while on patient rounds.

### Limitations

Limitations of study were that all possible error-prone abbreviations were not included, the post intervention study duration was limited to three months and sustainability could not be checked beyond that.

### Conclusion

‘Never Use’ lists are effective in reducing incidents of common error-prone abbreviations, discipline wise variation is observed and adoption of such lists is highly recommended. However, the lists should be comprehensive, regularly updated and educational interventions should be more comprehensive with integration into patient medical charts and pocket friendly flash cards may also be provided for better outcomes. Enforcing a policy to prohibit the use of EPAs while prescribing will also be helpful. The study adds to the research domain by establishing the impact of never use list of EPAs and opens avenues for further research on more comprehensive intervention and a policy inhibiting use of error prone abbreviations.

## Data Availability

No data is available

## Acknowledgments

Nil

## Other Required Statements

The article is SQUIRE compliant.

